# “Another world of pain” – Athlete and Sport Physiotherapist perspectives on the unique experience of pain in sport

**DOI:** 10.1101/2023.12.28.23300487

**Authors:** Ciarán Purcell, Caoimhe Barry Walsh, Garett Van Oirschot, Brona M Fullen, Tomás Ward, Brian M Caulfield

## Abstract

**Objectives:** To explore athlete’s and sports physiotherapists’ experiences of sports-related pain in the upper and lower limb.

**Methods:** Using a constructivist and pragmatic perspective we carried out focus groups comprising a deliberate criterion sample of athletes and sports physiotherapists. We used a topic guide that moved from open exploratory questions to questions focusing on the phenomena of sports-related pain in athletes. We coded, developed candidate themes and refined finalised themes using reflexive thematic analysis. A member of our research team acted as a critical friend adding additional perspectives. We followed the Consolidated Criteria for Reporting Qualitative Research (COREQ).

**Results:** We completed five focus groups comprising twelve athletes (n=5 female, n=7 male) and four sports physiotherapists (n=4 male) including one initial pilot (two athletes). We developed four final themes (I-IV) and nine subthemes (a-i). I) Athlete Pain Lens ( a - pain is part of being an athlete, b - pain shapes the life of an athlete) II) Exploring And Navigating Pain (c- the sports-related pain spectrum, d- making sense of pain) III) The Emotional Toll of Pain (e - challenging emotions, f - the impact of time) IV) Coping, Community and Communication (g - coping with Pain, h - influence of community and support network, i - communication-the broken key).

**Conclusion:** We highlighted the distinct and challenging phenomenon of sports-related pain experienced by athletes and physiotherapists. Through effective communication members of the athlete’s community may recognize, and adjust to these challenges.

## Introduction

With the rise in the commodification of athletes in elite sports and the focus on objective over subjective markers to quantify performance and health, it is more important now than ever before to listen to the voice of the athletes when they share their pain experiences before it is lost.^1^ Despite the focus on sports-related injury and availability to compete, a spotlight on sports-related pain has emerged.^2,3^ The International Olympic Committee (IOC) recommend a multidimensional approach to athlete pain that goes beyond traditional structure-based understanding to include aspects of pain perception and experience across five key domains: neurophysiological, biomechanical, affective, cognitive, and socioenvironmental.^4^ Psychosocial aspects such as history of stress and coping responses have been linked with injury rates.^5^ Furthermore, sports-specific sociocultural aspects such as attitudes and culture encouraging athletes to be “tough’’ and “stay in the game’’ have been linked to injury risk, injury rates and timely access to healthcare and treatment. The context of the pain experience has been highlighted as a key component of contemporary pain science, where the same pain experience may be interpreted as stressful or alarming in one context and as a helpful indicator to manage resources in another.^6^

Qualitative research can harness the athlete’s voice and explore the context of their experience which may ultimately lead to enhanced management strategies as has occurred in sports-related injury.^7^ Clinicians can feel underequipped when it comes to effectively understanding and assessing athlete pain.^8^ Traditionally, qualitative research has focused on the separate experiences of athletes and clinicians when it comes to pain, much of which has focused on low back pain.^9^ ^10^ To truly understand the sports-related pain phenomenon whilst preserving the voice of the athlete there is a need to gather both athletes’ and physiotherapists’ shared definitions and experiences.

We aimed to explore athlete and sports physiotherapists shared experiences and perspectives of upper and lower limb pain in a sporting context through mixed focus groups.

## Methods

We conducted a qualitative study with mixed focus groups combining athletes and sports physiotherapists and followed the consolidated criteria for reporting qualitative research (COREQ).^11^ Ethical permission was granted for our study by the UCD Human Research Ethics Committee. (LS- 22-40-Purcell-Caulfield)

### Positionality statement

As a sports physiotherapist and clinician my (CP) move to research was one inspired by pragmatism and constructivism drawing on the relevant knowledge from elements of empirico-analytical, interpretive and critical research paradigms.^12,13^ My motivation stems from investigating solutions to real-world problems and clinic queries where the best method is often the one that works. I recognise the need for hearing the patient’s (athlete’s) voice. My doubts, assumptions and beliefs drive my position of a value-driven axiology.^14^ I recognise that being a sports physiotherapist and PhD researcher influences discussions.^12^ I embrace my position to understand athletes’ and clinicians’ upper and lower limb pain experiences through taking a reflexive approach.^15^ Alongside eight years of clinical experience assessing and treating athletes, I (CP) have gained qualitative research experience through an MSc in Sports Physiotherapy. GvO (moderator) is a PhD candidate and sports physiotherapist with seventeen years of experience. BC (neutral observer), is a Professor in physiotherapy with twenty years of research experience. CBW, (critical friend) is a physiotherapist and PhD researcher in the field of chronic conditions with experience in the conduct of focus groups.

### Participants

We recruited a deliberate criterion sample. We sought athletes spread across multiple sports of varying ages, genders and competition levels, with experience of an upper or lower limb pain episode requiring sports physiotherapist assessment within the last year. We recruited sports physiotherapists with three or more years of experience currently working with athletes weekly from a range of clinical and sports settings. The notion of data saturation has been contested given that data and themes are generated through interpretation and there can be no objective point at which no new themes “emerge” in an active constructivist approach.^16^ Therefore, we ceased sampling when sufficient depth of information on sports-related pain experience was gathered. We recruited athletes through university sports clubs and local sports clubs’ email and communication channels. We contacted sports physiotherapists registered with the Irish Society of Chartered Physiotherapists Sports and Exercise Medicine Group (CPSEM) working across a variety of sports and settings email channels and networks. We offered participants their preference of either a fully face-to-face or fully online (zoom) focus group.

### Protocol

We sent participants a pre-participation questionnaire to capture background demographics, sporting context and pain history. We collected information from physiotherapists relating to their experience working in and participating in sports and any additional sports physiotherapy-related qualifications. We used a step-by-step approach which included: brainstorming, ordering, timing and phrasing of questions, obtaining feedback from peers, revising and piloting to develop the topic guide for the focus groups.^17^ The topic guide moved from broad introductions and context to more focused questions concerning athlete pain definitions, understandings and experiences.(Appendix A) Two hours were allocated to cover all questions which were open and encouraged interaction between participants. We altered the sequencing of questions between focus groups in line with an iterative and reflexive approach.^18^ We commenced each focus with ground rules focusing on confidentiality, equity and respect for diverse views and opinions and we requested that athletes share their opinions before physiotherapists to moderate the power imbalance. We endeavoured to separate athletes and physiotherapists who previously worked together. We completed audio recordings of all focus groups to facilitate transcription.

### Data Analysis

We transcribed the focus groups verbatim and published the uncoded and anonymised dataset in an online repository. (10.17632/t47tw94mzd.) Using a thematic analysis approach we began our data analysis following the initial pilot focus group informing sampling, data collection and analysis for subsequent focus groups in an iterative manner. I, (CP) read the transcripts in their entirety, added initial observations and notes and subsequently carried out initial coding of the entire transcripts (both semantic and latent). One-third of the transcripts were given to CBW to code independently. We discussed, compared and refined initial codes using a critical friend approach to incorporate breadth and variation in perspectives.^19^ CP then reread transcripts and updated coding where necessary before codes were compared, with similar codes being highlighted and grouped to form rough clusters of candidate themes. The team reviewed candidate themes in the context of the research and considered the data available to support each theme. We merged some themes and adjusted others at this refinement stage. We applied titles and developed a final thematic map.

### Patient & Public Involvement

A convenience group of five athletes, (female: n=3, male: n=2) from a range of competition levels and sports formed a PPI panel for this research project. They reviewed and commented on the research questions, topic guide and information provided to participants before the focus groups ensuring adequate athlete representation and appropriate participant burden. Their feedback was incorporated into the design and delivery of the final focus groups and their input was sought in the presentation of the final results and the inclusion of the final results in an initial Delphi survey which will inform the development of a set of athlete pain assessment guidelines.

### Equality, Diversity & Inclusion

Participants across gender, age, competition level and sport were sought for this study, our purposive sampling included local clubs both within and outside of the University setting facilitating diversity of the sample from an age and socioeconomic demographic perspective. We offered face-to-face as well as Zoom focus group options to increase accessibility. Our research team span a range of experiences and backgrounds.

### Results & Discussion

Five of the athletes had a background in physiotherapy practice (studying or practising physiotherapy across various settings). All physiotherapists were either current or previous athletes. We assigned athletes and physiotherapists alphanumeric participant IDs beginning with the letter “A” or ‘’P” respectively. Participant IDs were known only to CP, the lead researcher. Once participant IDs were allocated and analysis was complete all records of participant information were deleted ensuring anonymity. Athletes and physiotherapists discussed and explored shared experiences and concepts and so we merged the data for coding and analysis purposes. We identified codes in (i) athletes only, (ii) physiotherapists only and (iii) both athletes and physiotherapists. Figure 1 displays the final themes and the corresponding codes assigned to each theme. Figure 2 is a thematic map of the themes and subthemes relevant to sports-related pain.

**Table 1.**
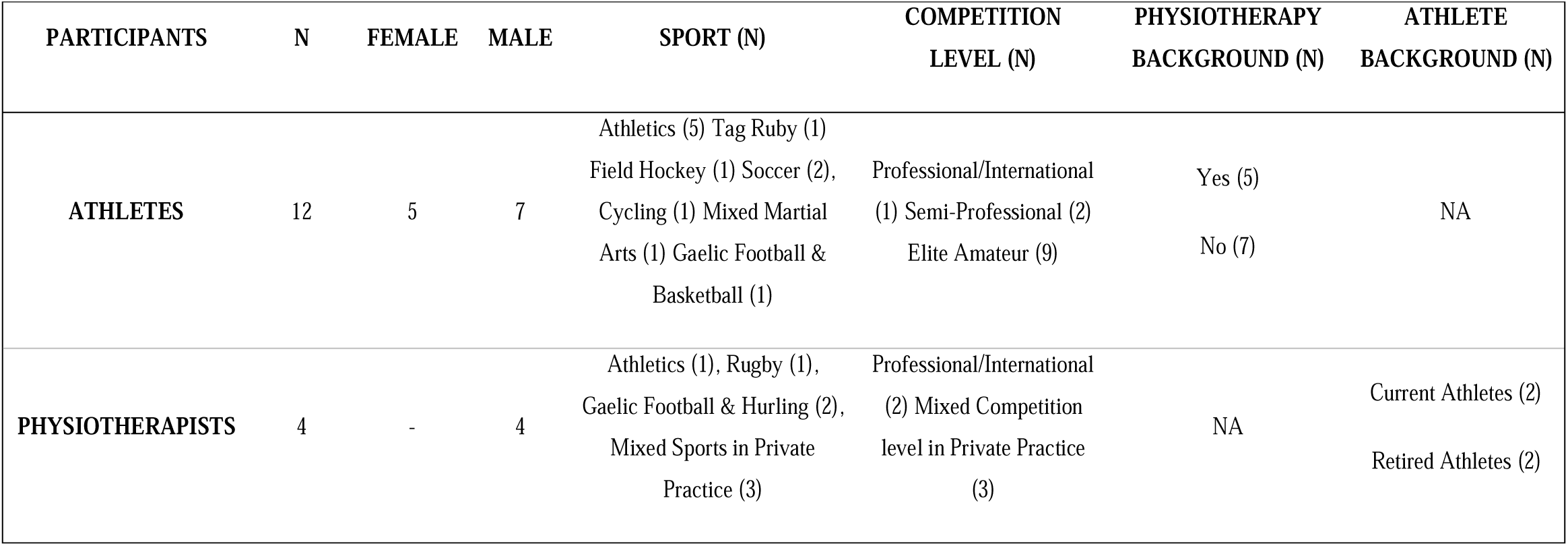
Participant Demographics.

| PARTICIPANTS | N | FEMALE | MALE | SPORT (N) | COMPETITION<br>LEVEL (N) | PHYSIOTHERAPY<br>BACKGROUND (N) | ATHLETE<br>BACKGROUND (N) |
| --- | --- | --- | --- | --- | --- | --- | --- |
| ATHLETES | 12 | 5 | 7 | Athletics (5) Tag Ruby (1)<br>Field Hockey (1) Soccer (2),<br>Cycling (1) Mixed Martial<br>Arts (1) Gaelic Football &<br>Basketball (1) | Professional/International<br>(1) Semi-Professional (2)<br>Elite Amateur (9) | Yes (5)<br>No (7) | NA |
| PHYSIOTHERAPISTS | 4 | - | 4 | Athletics (1), Rugby (1),<br>Gaelic Football & Hurling (2),<br>Mixed Sports in Private<br>Practice (3) | Professional/International<br>(2) Mixed Competition<br>level in Private Practice<br>(3) | NA | Current Athletes (2)<br>Retired Athletes (2) |

| PARTICIPANTS | N | FEMALE | MALE | SPORT (N) | COMPETITION<br>LEVEL (N) | PHYSIOTHERAPY<br>BACKGROUND (N) | ATHLETE<br>BACKGROUND (N) |
| --- | --- | --- | --- | --- | --- | --- | --- |
| ATHLETES | 12 | 5 | 7 | Athletics (5) Tag Ruby (1)<br>Field Hockey (1) Soccer (2),<br>Cycling (1) Mixed Martial<br>Arts (1) Gaelic Football &<br>Basketball (1) | Professional/International<br>(1) Semi-Professional (2)<br>Elite Amateur (9) | Yes (5)<br>No (7) | NA |
| PHYSIOTHERAPISTS | 4 | - | 4 | Athletics (1), Rugby (1),<br>Gaelic Football & Hurling (2),<br>Mixed Sports in Private<br>Practice (3) | Professional/International<br>(2) Mixed Competition<br>level in Private Practice<br>(3) | NA | Current Athletes (2)<br>Retired Athletes (2) |

**Figure 1.**
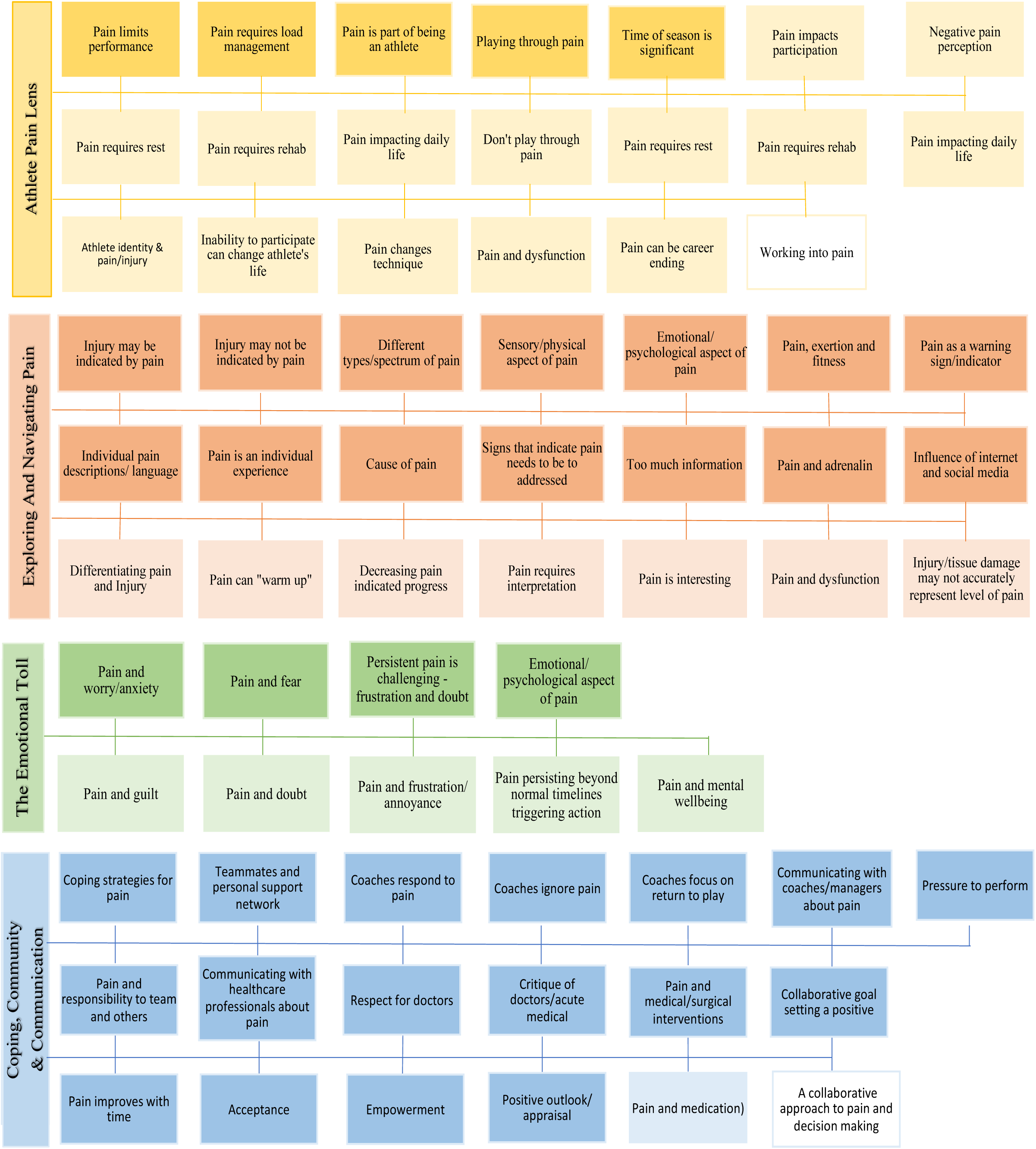
Themes (Athlete Pain Lens; Exploring & Navigating Pain; The Emotional Toll; Coping, Community & Communication) and codes. Dark shading – indicates codes that were present in athletes and physiotherapists. Light shading – indicates codes that were present in athletes only, No shading – indicates codes that were present in physiotherapists only

**Figure 2.**
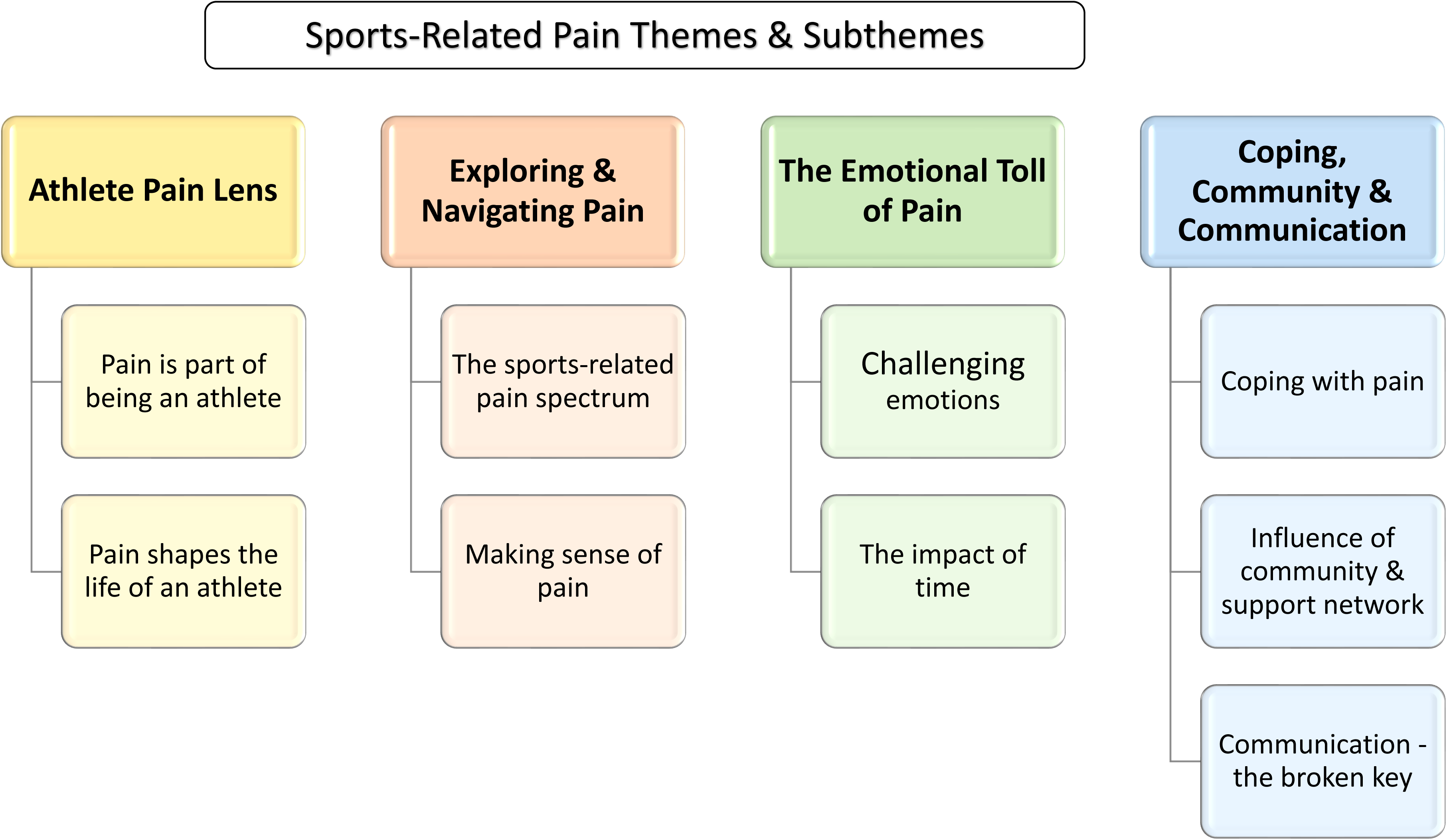
Athlete pain experience themes and subthemes. The themes for each part of this series represent a row in the priorities for pain assessment pyramid, this paper presents the bottom row, or foundation of the pyramid. This comprises the four themes that describe the “athlete pain” experience alongside the constituent subthemes. The middle row contains the three themes that describe the “athlete pain assessment” experience that will be presented in part two of this series. In the top row, the “priorities for future athlete pain assessment” themes and subthemes are to be presented in part three and will therefore build on the themes from the previous two papers.

#### Theme 1. The Athlete Pain Lens

Theme 1 includes two subthemes; 1.1 – pain is part of being an athlete and 1.2 – pain impacts and shapes the life of an athlete.

***1.1 Pain is part of being an athlete***

Participants shared how pain is an inherent aspect of being an athlete, shaping their identity. The interaction between pain, exertion and fitness is an enmeshed aspect of sports-related pain experience with athletes acknowledging that pain or muscle soreness can be a sign of effort towards the attainment of fitness. In line with contemporary research, playing through pain whether it be exertion-related or otherwise was commonplace behaviour reported by both sports physiotherapists and athletes albeit with potentially negative consequences.^20^ The main concern athletes had was the impact pain has on their performance, particularly during competitive parts of the season.

Physiotherapists also acknowledged the impact the time of the season has on pain behaviours.

> *“I find that you live with pain. It’s not that you wait for the day for pain to come or go.’’*

> – A01

> *“I’m happy to run through pain all summer and just my only worry is that it impacts*

> *performance.”* – A08.

> *“Because depending on time of season, what’s coming up, .. changes someone’s, maybe not how they experience pain but certainly how far they’re willing to take it.” – P04*

***1.2 Pain impacts and shapes the life of an athlete***

Athletes and Physiotherapists alike defined pain as something that influences an athlete’s ability to participate and perform in their sport. Athletes described how pain also impacts their daily life outside of sports with the combination of the physical and emotional experience having the potential to influence their identity. Previous research has documented how the impact of pain on daily life and mental wellness can force athletes to contemplate their future in sport, something one athlete in this study resonated with.^21^

> *“I suppose physical pain can often translate into being an emotional pain, not being able to participate can change the life of an athlete.”* – A04

> *“It happens me all day, every day, there’s times that I limp because of it and I can’t walk. I’ve had to adjust my lifestyle. I can’t wear certain shoes, I can’t just ..run for a bus I need a good long*

> *warm up and I am meant to be an athlete.”* – A08

> *“I’m able to run through it but it causes me so much pain that I contemplate retiring.”* – A08

#### Theme 2. Exploring And Navigating Pain

Theme 2 includes two subthemes; 2.1 – the sports-related pain spectrum and 2.2 – making sense of pain. Athletes experience a variety of unique pain encounters. They interpret and learn about their pain to determine whether each pain episode is part of the normal pain experience, or if it is associated with potential injury and subsequently warrants action.^20,22^ Pain is neither predictable nor repeatable. The abundance of available information through the internet and various sources is often more of a hindrance than a help. Athletes appear to have more competition for their attention than previous generations, as discussed by coaches of Generation Z athletes.^23^ All of these aspects contribute to a complex phenomenon that is not easily understood and warrants greater attention, appreciation and representation.

***2.1 The sports-related pain spectrum***

In line with contemporary research, pain experience existed along a spectrum for athletes and physiotherapists alike who described a variety of pain presentations from routine training-related muscle soreness, to “niggles”, to pain that is more sinister and may be a cause for concern.^24^ Pain perception has both sensory and emotional aspects that, participants described, are influenced by a multitude of sports-related activities from a routine warm-up to the rush of adrenalin and intense focus experienced when competing.^22^ This situational and contextual pain perception served the purpose of deferring the pain experience and facilitating athletes’ performance during competition.

> *“It kind of has physical implications and psychological as well yeah”.* – A11

> *“For the majority of anything from a niggle to an injury, if you warm up you can still run on*

> *it.” – A01*

> *“The adrenaline of it all takes over I’m able to finish the rep or finish the race but as soon as I stepped off the track, I said to my coach on both occasions like ah somethings gone wrong.”*–

> A08

***2.2 Making sense of pain***

Differentiating between pain and injury is a challenge athletes and physiotherapists often grapple with. In many cases, injuries may be indicated by pain whilst in others it is not. The amount and intensity of pain an athlete perceives are influenced factors beyond tissue damage, something which athletes and physiotherapists found frustrating to reconcile.^4^

> *“That is something I definitely struggle with from an athlete perspective is when does pain become something that stops you from competing…as opposed to finding that place where you know*

> *it’s safe to continue.”* – A12

> *“I don’t think it relates to pain, I don’t think pain relates to level of injury, I think it’s a very loose connection to make, especially an acute injury.”* – P04

Exploring the meaning of pain, athletes and physiotherapists acknowledged the individual nature of pain enlisting individual pain descriptions and language to describe their experiences. Accurately representing pain required consideration and time for reflection with successful communication hinging on the physiotherapist understanding and valuing the athlete’s unique lexicon.

> *“I think everyone kind of expresses themselves differently.”* – A11

> *“I suppose as long as your physio understands your language, that’s the key thing and that’s where the relationship with your physio is actually kind of important.”* – P03

Contributing to feelings of frustration, athletes are also faced with an abundance of information from different sources. The availability of large swathes of information makes it difficult to identify the most important elements and operating in a contemporary world adds additional complexity^23^ Athletes and physiotherapists discussed how the internet, social media and data derived from wearable technologies might provide a source of helpful information but often exacerbates the dilemma of deciphering pain due to the nature of comparison on social media and the abundance of non-evidence based information, particularly for athletes who have poorer levels of digital health literacy.

> *“I think people get really confused about the whole situation, they’ve read stuff, they’ve probably been to other practitioners before, they’ve gone around the mill so they can be a little bit of a challenge in terms of figuring out where they are from both a pain point of view, function point of*

> *view.”* – P03

> *“Like Instagram and people influencing you and you know like I was changing exercises every single day trying to solve my pains … the stress and the thought process.”* – A07

Despite the challenges, athletes valued the protective role pain plays and were intrigued to learn more both about, and from, their pain. Pain acted as a warning sign or indicator that athletes and physiotherapists used when deciding which pain and load management strategies to apply.

> *“But for me, it’s kind of a feedback mechanism for the injuries to kind of guide me on what I can and can’t do. If the body signals to kind of back off on things and try to kind of use it as a guide*

> *or to teach you how far to push things with injuries and stuff.”* – A09

#### Theme 3. The Emotional Toll Of Pain

Theme 3 includes two subthemes; 3.1 – challenging emotions and 3.2 – the impact of time on the emotional aspects of pain. Athletes experience a range of emotions expressing the challenges and psychological toll of navigating pain.^22^ Guilt, worry, fear, frustration, and doubt are part of the pain experience. These emotions were also observed in sports-related pain in this study and were related to an athlete’s inability to participate and perform in their given sport and indeed daily life activities.^25^ These emotions tended to progress insidiously, with time highlighting and heightening the psychological aspects of pain.^20^

**3.1 Challenging emotions**

Athletes reported they often feel worried and anxious owing to the lack of certainty of the potential causes, consequences and outlook of pain. Pain persisting beyond expected timeframes encroached on everyday life and heightened concerns. Pain and feelings of doubt negatively impacted athletes’ capacity to make sense of and manage their pain effectively. Fear, doubt and frustration were often synonymous with athletes’ experiences of pain and emotions have previously been found to impact an athlete’s ability to maintain activity levels when injured.^26^ Athletes spoke about how they fear pain intensifying and are acutely aware of the impending impact it may pose on their ability to participate in sport. At times, the inertia accompanied by this cocktail of emotions left athletes feeling guilty for not being more proactive and blaming themselves for their situation.

> *“The fear that it’s going to get worse or feeling like it’s going to stop me from playing.”* – A02

> *“When you go through the pain for that long you want it to not be there you start to not trust your own perception”* – A03

> *“But then I do kind of feel bad because if it’s something that happened before, then I know I should have been doing my rehab .. so it’s kind of my fault.”* – A01

**3.2 The impact of time on the emotional aspects of pain**

The trial-and-error nature of pain assessment and management was often at odds with the protracted timelines athletes have to make decisions and be ready for their next competition, leading to frustration. Participants described how repeated pressurised decisions contribute to the cumulative emotional toll which negatively impacts an athlete’s physical and mental well-being, reflecting research findings. ^21^ Athletes discussed the negative impact of pain on their sleep, contributing to fatigue and further fuelling a negative pain and quality of life cycle.

> *“Obviously when pain would reach a point that it takes you out of the field of play it can really affect you mentally and it’s difficult to deal with and it can affect all walks of life just away from the sport”* – A05

> *“Yes it was really frustrating because it was like a circle of not being able to sleep and that kind of causing disruption ..to feel just tired and generally fatigued throughout the day and then fairly anxious and depressed that you can’t train”* – A10

#### Theme 4. Coping, Community and Communication

Theme 4 includes three subthemes; 4.1 – coping with pain 4.2 – influence of community and support networks and 4.3 – communication, the broken key.

***4.1 Coping with pain***

Applying transferable skills acquired in their sports, athletes discussed using a variety of coping strategies including managing their attention and maintaining their fitness with non pain-provoking cross-training modalities. Athletes can use the additional time to reflect and foster other interests and hobbies away from their sport which may help manage stress, an important aspect of pain management.^5^ Athletes acknowledged the often-unrelenting nature of the pursuit of excellence within sport and the opportunities afforded during pain and injury.

> *“Sometimes you just want to ignore pain and just get through it or whatever.” - A05*

> *“Just kind of did my own thing, kind of eased off..worked around it because I do kind of a broad range of stuff like and almost always work around injuries” – A09*

> *“You have other parts of your life that you can focus on like you’re able to focus on different types of training .. and like for example I really enjoy gardening so like whenever I’m injured, I tend to go and garden more because well I can’t train but also it’s a really nice outlet for me to relieve some of that stress.” – A08*

***4.2 Influence of community and support networks***

Understanding and managing pain can be a solitary pursuit however athletes identified that they are part of a wider community. Relationships with friends, family, partners, teammates, colleagues, coaches and healthcare practitioners play a pivotal role in athletes understanding, managing and coping with pain.^27^ Emotional support for athletes came from personal relationships and teammates who were often the most appropriately positioned to offer context-specific empathy and advice.

Conversely, athletes involved in individual sports were acutely aware of the void of comradery.

> *“My current partner .. she was just like .. go back and get some sleep and take care of yourself for a few weeks.” – A10*

> *“The players we all live together they are the ones who would probably check in on you more in that aspect .. asking you from the perspective of how you are feeling about it.” – A05*

> *“I think it’s better that if you’re in a team sport, you could talk to some of your teammates like A02 could. Like it’s sometimes hard on your own.” – A01*

Athletes described how they spend a significant amount of time with coaches with relationships developing over years in many cases. The support an athlete feels from their coach is inextricably linked to the coach’s attitude to pain.^25^ Participants discussed how some coaches tend to ignore pain, focusing on return to play whilst others respond to pain acknowledging the impact it has on the athlete and the strategies needed to best manage it. Physiotherapists acknowledged the integral role coaching staff play in managing athlete pain, investing time in explaining the complexities behind return-to-play timelines and the variable nature of pain that is unique to each athlete.

> *“The coaches very rarely ever asked about how are you getting on, how are you dealing with it, it was always just when are you back. So, they don’t tend to look at you as much as a person and*

> *ask you about how you are getting on with it and how are you feeling about it.”* – A05

> *“Because in different scenarios management play a huge role in it like is there somebody that is going to look and say you know, we’ll sit you out this game or you don’t have to train Wednesday*

> *but you’re still playing Saturday or trying to get a balance.”* – A07

> *“I find I spend a lot of time actually educating managers on injuries and the likely timeframes and how that might change depending on how they’re doing.”* – P02

***4.3 Communication – the broken key***

Central to successful and supportive relationships is effective communication regarding the experience of pain.^27^ Physiotherapists and athletes spoke about how they work together and with other members of an interdisciplinary team at various stages throughout the pain journey. Adequately reporting and describing pain to clinicians was often challenging for athletes owing to the time constraints of clinical and sports-based assessments. Both athletes and physiotherapists described how frustration can build in persisting pain presentations or when the cause of pain is unclear.

> *“Because I, actually, before I go into a physio, I do write down what I’m going to say, because more often than not I’d leave forgetting to mention things and you’re like, oh god, well it’s*

> *gone now and I’m never going to say anything.”* – A11

Athletes and physiotherapists discussed how physicians often sit at the top of the decision-making tree when it comes to managing athlete pain medically. As a result, some athletes find the decisive nature of these clinical decision making interactions refreshing, providing clarity. In stark contrast, athletes can also find interactions with physicians limiting and reductionist, with physiotherapists also echoing this frustration.

> *“Whenever I feel pain, I go to the Doctor and they confirm what my pain was and I’m reassured that what I felt was real and then I follow the rehab protocol.”* – A08

> *“That’s very hard .. yes this pain like that thing in my foot I’m like I’ve had this years ..how do you get a sports doc to see that beyond just the MRI and what it shows you?’’* – A08

> *“With the doctor instead of constantly saying to everyone we’ll just MRI your back maybe they start to get a bit better too at knowing who they should be referring for medical imaging because*

> *we’re picking up the bloody pieces after it..”* – P02

Athletes shared how in sport, the pressure to perform and rapidly return to play, a deep sense of responsibility to teammates and playing through pain culture can be at odds with the time and space needed to effectively communicate the pain experience within their support network. Physiotherapists also noted the effect the team dynamic of sport has on an athlete negotiating their pain.

> *“The team environment and how competitive it was at the time, you couldn’t really say to the management unless it got to a point where like there’s fairly you know something sinister or*

> *something”* – A07.

> *“There is a little bit of a sense of community, teammates and stuff that’s going on as well where ‘who am I letting down?’ ..so you know.”* – P03

When communication and the ensuing support are hampered, athletes found that their resources for coping are reduced and the likelihood of seeking medical and surgical interventions can increase.

Conversely, when communication is effective, collaborative decision-making can occur with comprehensive pain management plans being devised. This was often accompanied by a more positive outlook with athletes reporting feelings of acceptance and empowerment as key markers of moving forward and coping with pain.

> *“Until you kind of accept it and then you sort of say to yourself okay right, what’s my path to recovery? I think you’re at nothing with most injuries to be honest like you have to have that level of*

> *acceptance.”* – A09

Athletes resourcefully employ a range of strategies to improve their understanding and manage their pain. An athlete’s personal, sporting, and medical community forms an ecosystem which offers additional strategies beyond those available on an individual level. Accessing these strategies often hinges on effective communication between the athlete and their support network.^28^ The importance of communication between athletes, their coaching staff and clinicians is well documented and impacts performance and injury rates as well as the athlete pain experience.^29^

### Clinical & Research Implications

Sports-related pain is a complex experience unique to each athlete that requires interpretation and can be challenging to understand, cope with and manage. All members of the wider athlete support network (including coaching, sports science and sports medicine) must appreciate the significant impact of pain on performance and quality of life and recognise the steps we can all take to help athletes make sense of their pain and manage it more effectively This includes appreciating the spectrum of contextual factors presented in this paper. There is scope for future work to explore the sports-related pain assessment experiences and priorities of both athletes and physiotherapists to improve assessment and indeed treatment and management selection.

### Limitations

Whilst every effort was made to collect experiences from a diverse range of athletes and physiotherapists and variety was achieved in sport, competition level and practice setting, the experiences gathered from these focus groups may not apply to all athlete pain settings. Notably, participants were all recruited from Ireland and whilst some of the female athletes also had a physiotherapy background no female sports physiotherapists were available to participate.

### Conclusion

This paper presents four key themes (Athlete Pain Lens; Exploring and Navigating Pain; The Emotional Toll; Coping, Community and Communication), capturing the unique entity of athlete pain experience through combined focused groups of athletes and sports physiotherapists.

### What is already known on this topic?

- Pain is a complex biopsychosocial experience that is prevalent in athlete’s influencing performance and quality of life

### What this study adds

- Sports-related pain is unique to each athlete forming part of their identity.

Sport-related pain poses challenges for athletes and Physiotherapists to interpret, diagnose and manage and is influenced by a range of emotions and contextual factors that can be addressed in a comprehensive biopsychosocial model of pain assessment and management.

### How this study might affect research practice or policy

- Clinicians and athlete support staff can consider the range of internal (emotional, cognitive) and external (socioenvironmental and support networks) contextual factors influencing the experience of pain in athletes and appreciate the challenges this and the influence effective communication can have. There is scope for future research to explore the unique nature of the athlete pain assessment

## Data Availability

All data produced are available online at:10.17632/t47tw94mzd.1 https://data.mendeley.com/datasets/t47tw94mzd/2

## Acknowledgements

A huge thank you to Alison Keogh and Aisling Lacey for their consultation and expertise in qualitative research methods in sport and exercise.

