## Appendices for "“Another world of pain” – Athlete and Sport Physiotherapist perspectives on the unique experience of pain in sport"

**Appendix A Focus Group Topic Guide/Question Route**

*First 10-15 mins. Ground rules (equal voices, respect opinions confidential nature of session) Participants are invited to introduce themselves, say a little about themselves*

**Question 1) What does pain mean to you in terms of being an athlete?**

Prompts:

- Are there different types of pain?
- Injury vs training soreness?
- Do you think pain means different things to different athletes?
- Is short-term pain different to long-term pain?

**Question 2) Can you recall an upper or lower limb pain experience you had in the last year?**

Prompts:

- Was it an acute pain episode or recurrent/lingering?
- What were the physical feelings/sensations associated with that pain?
- What were the emotions and thoughts associated with that pain?
- When was it, what time of the year/season? How did it change your day-to-day and sporting activities? How did you manage/address this pain?
- Physiotherapists - can you share a personal upper or lower limb pain experience or that of an athlete you worked with?
